# Prevalence and predictors of virological failure and quality of life of people with HIV/AIDS at a Municipal Hospital, Volta Region -Ghana

**DOI:** 10.1101/2025.08.01.25332761

**Authors:** Kwaku Gyimah Peprah, Faith Agbozo, Mavis Pearl Kwabla, Worlanyo Tashie, Joyce Berkumwin Der

## Abstract

**Background:** Despite several interventions to eradicate HIV/AIDS globally, virological failure continues to threaten the goals of anti-retroviral therapies (ART) and quality of life (QoL) of people with HIV/AIDS (PWHA). This study assessed the prevalence and predictors of virological failure and QoL of PWHA.

**Methods:** A cross-sectional study was conducted at the ART clinic of a Municipal Hospital, from June to August 2023, to assess the socio-demographic, clinical/ medical data, and QoL of PWHA receiving therapy at the clinic. Participants were randomly selected and interviewed: their weight and height were taken and their clinic folders examined to assess virological failure status. Bivariate and multiple logistic regression analysis were conducted to determine the predictors of virological failure. Also, multiple linear regression was conducted to determine factors contributing to QoL of study participants.

**Results:** A total of 398 participants comprising of 328 (82.41%) females, and with a mean age of 48.2 years (SD ± 11.71 years), were recruited into the study. The prevalence of virological failure was 6.03%. Factors such as forgetting to take ART (AOR = 2.87, 95% CI = 1.02, 7.51; *p*=0.04), being classified as baseline WHO staging II (AOR = 6.20, 95% CI = 1.91, 20.04; *p*=0.002), and HIV stigmatization (AOR = 3.97, 95% C.I. = 1.1, 14.25; *p*=0.035) were associated with virological failure. The overall QoL was good (75.35%). Having no comorbidities (R^2^=-2.7, *p*<0.0001), having social support (R^2^= 3.94, *p*<0.0001) and receiving an average monthly income (R^2^=2.03, *p*=0.002) contributed to good QoL.

**Conclusion:** Virological failure in the municipality exceeded the 5.0% target set by the Joint United Nations Programme, despite majority of the study participants presenting with good QoL. The National AIDS Control Programme should consider long-acting injectable therapy for PWHA struggling to adhere to medication.

## Introduction

The Human Immunodeficiency Virus/Acquired Immune Deficiency Syndrome (HIV/AIDS) menace still remains a public health problem [1, 2]. Approximately 37.7 million individuals are infected with HIV/AIDS globally, with about 25.7 million cases occurring in sub-Saharan Africa [3]. In Ghana, about 334,095 people were estimated to be living with HIV/AIDS in the year 2023 [4]. Several interventions have been implemented in healthcare systems to reduce the burden of HIV/AIDS and subsequently control the HIV epidemic [1, 5–7]. Despite these interventions, virological failure continues to impede the health and quality of life (QoL) of people with HIV/AIDS (PWHA) [8].

Virological failure is a type of HIV treatment failure where the viral load of a person with HIV/AIDS fails to fall below 1000 copies/mL despite taking ART for at least 6 months [9, 10]. This treatment failure contributes to high HIV mortality, drug resistance and poor QoL among PWHA [11]. QoL is the feeling of overall life satisfaction, as determined by the mentally alert individual whose life is being evaluated, and it reviews the physical health, social support, financial security, spiritual and emotional well-being of these individuals [12]. QoL is an excellent prognostic indicator for diseases and its application can be used to support decision making which are needed to improve the health of PWHA [13]. Virological failure worsens the health of PWHA by increasing their viral load and reducing the body’s immunity [14]. The reduction in immunity facilitates the development of opportunistic infection, drug resistance and other psychiatric disorders such as depression and anxiety [15]. These worsens the QoL of PWHA [16].

Although the prevalence of virological failure has been determined across several countries in Africa [11, 17], only few studies have achieved 95% viral suppression rate [18] as established by the Joint United Nations Programme on HIV/AIDS (UNAIDS). In Ghana, several studies [19–22] suggest that virological failure continues to impede the goals of ART and UNAIDS.

Ketu-South municipality is among the municipalities in the Volta Region with a high HIV/AIDS burden, noted for risky sexual behaviors such as prostitution [23]. Despite this assertion, there appears to be paucity of data on virological failure and QoL of PWHA on ART in the municipality. Few researchers have conducted similar works in other municipalities [20, 24, 25], however, the social, cultural and behavior differences existing among communities and the need to strengthen HIV interventions in the municipality warrant the need to conduct this study. Therefore, this study aimed to determine the prevalence and predictors of virological failure and QoL of PWHA in the Ketu South Municipality.

## Methods

### Study design and setting

A hospital-based cross-sectional study was conducted in the Ketu South Municipality of the Volta Region of Ghana from 6^th^ June 2023 to 31^st^ August 2023. The Ketu South Municipality has a total land area of about 261 square kilometers, with a population of 253,122 as reported by the 2021 national population and housing census [26], and shares border with the Republic of Togo. Despite the municipality’s population being composed of farmers and traders, there is also the presence of high-risk behaviors, such as prostitution, which is commonly observed among border towns [27, 28]. Participants were recruited from the ART clinic of the Municipal Hospital, which serves as the major referral hospital for neighboring health facilities.

### Study participants

HIV-infected adults receiving therapy at the ART clinic were randomly recruited for the study. Participants were individuals who are 18 years old and above, and had been on ART for at least six months.

### Eligibility criteria

HIV-infected adults aged 18 years and above who had been on ART for at least 6 months, and had at least one viral load measured were recruited. Individuals who did not have complete medical records prior to the study were excluded. Also, individuals who did not give written informed consent were excluded from the study.

### Sample size calculation

The Cochran sample size formula was used to calculate the sample size for this study. With 95% confidence level, 5% sample error, 1.96 standard error and a prevalence of virological failure at 38% [29], the sample size for this study was computed. A 10% non-response rate was added to the calculated sample size to give a total sample size of 398.

### Sampling

A simple random sampling technique was used to select participants from the HIV population who visited the ART clinic of the Municipal Hospital. This method of sampling gives equal chance of selection to individuals who visited the clinic. The lottery method of simple random sampling was employed in this study. During the sampling procedure, unique numbers were assigned to PWHA who visited on clinic days. These numbers were sealed in a paper and placed in a small bowl. The sealed papers were thoroughly mixed for one minute and randomly selected without replacement, until the expected daily sample size was achieved. The selected individuals were approached and those who consented were recruited into the study. The ART clinic operated twice in a week. In each day, an average of 50 clients were reviewed by the ART clinic. A total of 30 study participants were randomly recruited in each clinic day until the entire sample size of the study was obtained.

### Data collection procedure

Study participants who visited the ART clinic on clinic days were randomly selected into the study. In a face-to-face interview with each participant, data on their drug adherence, social support, stigma, behavioral attitude, clinical and socio-demographics were obtained. Also, each participant’s hospital folder and database were assessed for their viral load and other medical data. The hieght and weight of study participants were measured to determine the body mass index (BMI) of study participants. QoL of study participants were assessed by administering a structured tool (WHOQOL-HIV BREF scale) adopted from the World Health Organization (WHO). These data were collected electronically using the Kobo Collect sotware and exported into microsoft excel 2016 spreadsheet.

### Quality of life instrument

The WHOQOL-HIV BREF scale was used to assess the QoL of study participants. The tool comprised of 31 questions, categorized under 6 domains, which assessed the physical health, psychological health, level of independence, social relationships, environment and spirituality of PWHA. Each question had a five option likert-type response which reflects the opinions of participants. Scores were assigned in ascending order, ranging from option 1 (lowest score) to option 5 (highest score). In some few questions, the scores denoted for the options selected were reversely scored. The total score for each domain was computed by adding the individual scores obtained from the set of questions that fall under each domain. The total scores obtained for each domain was then divided by the total attainable score for that domain and then expressed as a percentage. The QoL scores obtained was further categorized as excellent (scores ranging from 80-100), good (60-79) and poor (less than 60), based on their percentage scores [30].

### Statistical analysis

The dependent variables in this study were virological failure and QoL, whereas the independent variables were the characteristics of socio-demographics, social support, baseline HIV information, stigma, adherence and behavioral attitude of HIV-infected adults. Descriptive analysis was performed to determine the mean, standard deviation, frequencies and percentages for the continuous variables. In the inferencial statistics, bivariate logistic regression was performed to determine predictor variables that were associated with virological failure. Predictor variables with *p*-value less than 0.2 were fitted into the multiple logistic regression analysis. In the multiple logistic regression, variables with *p*-value less than 0.05 were considered as significant predictors of virological failure. Multiple linear regression analysis was also performed to determine factors that were associated with QoL of participants. At all times, a *p-*value < 0.05 was considered as statistically significant. All analysis were performed using STATA (version 13).

### Ethical consideration

Approval for this study was obtained from the Ethics Review Committee of University of Health and Allied Sciences (UHAS-REC A.5 [1] 22-23). Permission was sought from the Municipal Hospital before accessing the facility for the study. No minors were recruited in this study. Written informed consent was obtained from each participant after the objective and rationale of the study had been explained to them.

## Results

### Socio-demographic characteristics of people with HIV/AIDS

A total of 398 people with HIV/AIDS were included in the study, of which 328 (82.41%) were females. Also, 120 (30.15%) participants had no formal education whereas 139 (34.92%) and 100 (25.13%) had completed basic and junior high school respectively. The majority of study participants (193, 48.54%) were married whilst 115 (28.89%) and 44 (11.06%) were widowed and single respectively (Table 1).

**Table 1:**
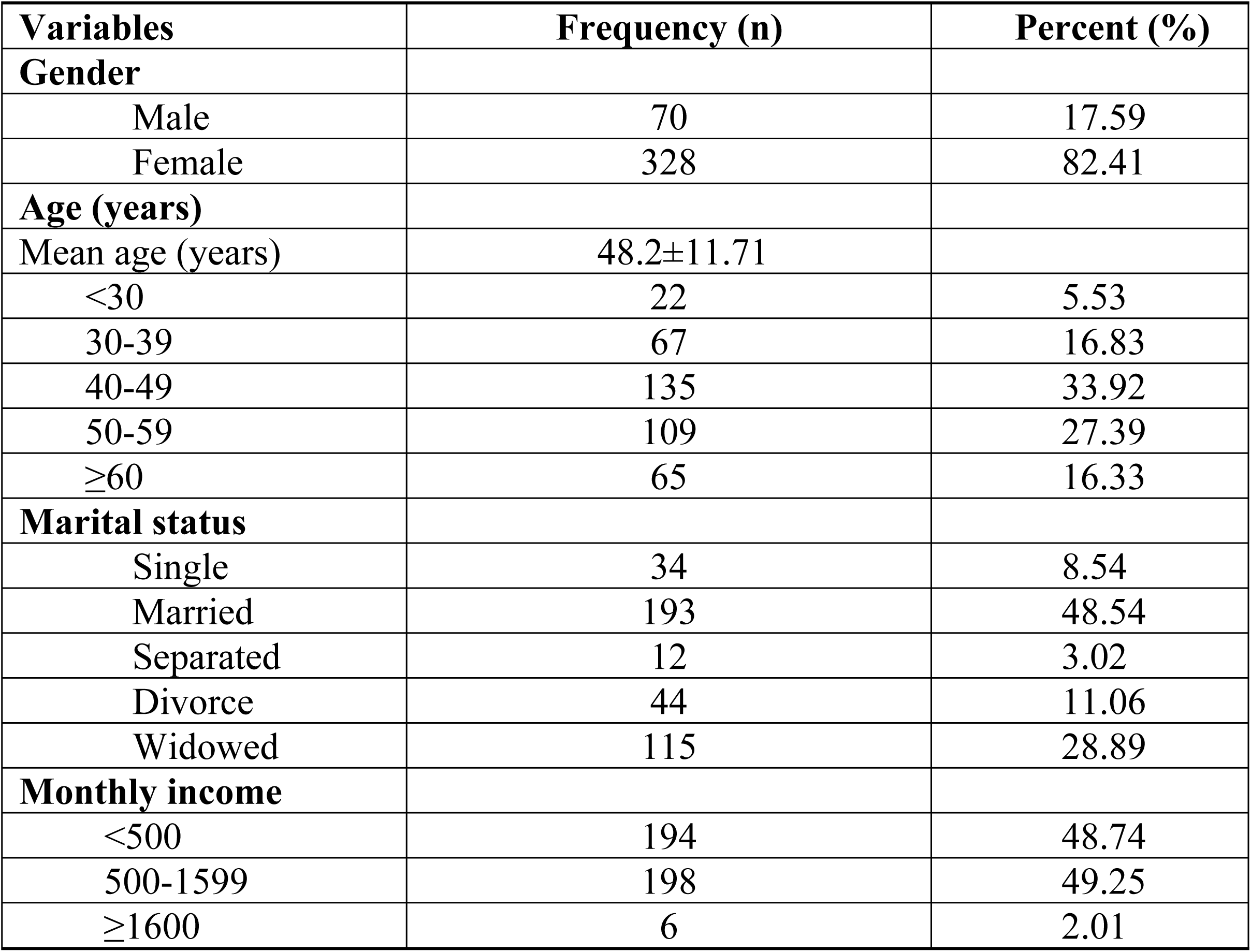
Socio-demographic characteristics of people with HIV/AIDS at a Municipal Hospital, Volta Region.

| Variables | Frequency (n) | Percent (%) |
| --- | --- | --- |
| **Gender** |  |  |
| Male | 70 | 17.59 |
| Female | 328 | 82.41 |
| **Age (years)** |  |  |
| Mean age (years) | 48.2±11.71 |  |
| <30 | 22 | 5.53 |
| 30-39 | 67 | 16.83 |
| 40-49 | 135 | 33.92 |
| 50-59 | 109 | 27.39 |
| ≥60 | 65 | 16.33 |
| **Marital status** |  |  |
| Single | 34 | 8.54 |
| Married | 193 | 48.54 |
| Separated | 12 | 3.02 |
| Divorce | 44 | 11.06 |
| Widowed | 115 | 28.89 |
| **Monthly income** |  |  |
| <500 | 194 | 48.74 |
| 500-1599 | 198 | 49.25 |
| ≥1600 | 6 | 2.01 |

### Clinical and adherence characteristics of people with HIV/AIDS

There were 88 (22.1%) study participants with comorbidities such as hypertension, tuberculosis and diabetes; 93 (23.37%) had a history of discontinuing their ART medication; and 262 (65.83%) had kept ART scheduled appointment. Also, 379 (95.23%) were currently taking TDF-3TC-DTG anti-retroviral combination therapy. Additionally, 108 (27.14%) participants had ever forgotten to take their ART medication. It was noted that 280 (70.35%) participants had their HIV status disclosed. Only 29 (7.34%) reported to have been stigmatized. This is summarized in Table 2.

**Table 2:** Clinical and adherence characteristics of people with HIV/AIDS at a Municipal Hospital, Volta Region.

| Variables | Frequency (n) | Percent (%) |
| --- | --- | --- |
| **Baseline WHO Staging** |  |  |
| Stage 1 | 330 | 82.91 |
| Stage 2 | 38 | 9.55 |
| Stage 3 | 28 | 7.04 |
| Stage 4 | 2 | 0.5 |
| **Final WHO Staging** |  |  |
| Stage1 | 388 | 97.49 |
| Stage 2 | 8 | 2.01 |
| Stage 3 | 2 | 0.5 |
| **Current ART Regimen** |  |  |
| AZT 3TC DTG | 19 | 4.77 |
| TDF 3TC DTG | 379 | 95.23 |
| **Presence of comorbid** |  |  |
| Yes | 88 | 22.11 |
| No | 310 | 77.89 |
| **Keeping scheduled appointment** |  |  |
| Yes | 262 | 65.83 |
| No | 136 | 34.17 |
| **Have you ever forgotten intake of ART** |  |  |
| Yes | 108 | 27.14 |
| No | 290 | 72.86 |
| **Forgotten ART intake during weekend** |  |  |
| Yes | 51 | 12.81 |
| No | 347 | 87.19 |
| **Social support** |  |  |
| Yes | 201 | 50.5 |
| No | 197 | 49.5 |
| **HIV disclosure** |  |  |
| Yes | 280 | 70.35 |
| No | 118 | 29.65 |
| **HIV stigma** |  |  |
| Yes | 29 | 7.29 |
| No | 369 | 92.71 |

AZT= Zidovudine; 3TC= Lamivudine; EFV= Efavirenz; NVP= Nevirapine; TDF= Tenofovir Disoproxil Fumarate; DTG= Dolutegravir; ART= Anti-Retroviral Therapy; WHO= World Health Organization.

### Factors associated with virological failure among people with HIV/AIDS

Table 3 summarizes the variables of participants that are associated with virological failure. People with HIV/AIDS who were grouped in baseline WHO stage 2 were 6 times more likely to develop virological failure compared with their counterparts in WHO stages 1 (AOR *=* 6.2; 95%CI *=* 1.91, 20.04; p = 0.002). Also, people with HIV/AIDS who forgot to take their ART medication were 2.78 times more likely to develop virological failure than those who always take their medication (AOR= 2.78; 95%CI = 1.02, 7.51; p= 0.04). People with HIV/AIDS who reported being stigmatized were 2 times more likely to develop virological failure than those who reported not being stigmatized (AOR = 2.11; 95%CI = 1.1, 14.25; p = 0.035).

**Table 3:** Factors associated with virological failure among people with HIV/AIDS at a Municipal Hospital, Volta Region.

| Variable | VF(%) | OR | *p*-value(95%CI) | AOR | *p*-value(95%CI) |
| --- | --- | --- | --- | --- | --- |
| **Age** |  |  |  |  |  |
| <30 | 3(12.51) | 1 |  |  |  |
| 30-39 | 8(33.33) | 0.86 | 0.83(0.21, 3.56) | 0.81 | 0.8(0.15, 4.16) |
| 40-49 | 5(20.83) | 0.24 | 0.06(0.54, 1.10) | 0.25 | 0.118(0.04, 1.41) |
| 50-59 | 6(25.0) | 0.37 | 0.18(0.08, 1.60) | 0.39 | 0.289(0.07, 2.18) |
| ≥60 | 2(8.33) | 0.2 | 0.09(0.03, 1.29) | 0.19 | 0.128(0.02, 1.6) |
| **BMI** |  |  |  |  |  |
| Underweight | 3(12.5) | 1 |  |  |  |
| Normal | 13(54.17) | 0.37 | 0.14(0.09, 1.42) | 0.34 | 0.183(0.07, 1.65) |
| Overweight | 8(33.33) | 0.5 | 0.34(0.12, 2.06) | 0.55 | 0.49(0.11, 2.93) |
| **Baseline WHO stage** |  |  |  |  |  |
| Stage 1 | 15(62.50) | 1 |  |  |  |
| Stage 2 | 6(25.0) | 3.94 | 0.08(1.43, 10.85) | 6.2 | 0.002(1.91, 20.04) |
| Stage 3 | 3(12.5) | 2.52 | 0.16(0.68, 9.29) | 2.12 | 0.337(0.45, 9.83) |
| **ART discontinuation** |  |  |  |  |  |
| No | 16(66.67) | 1 |  |  |  |
| Yes | 8(33.33) | 2.5 | 0.03(1.07, 5.84) | 2.08 | 0.156(0.75, 5.73) |
| **Forgetting ART intake** |  |  |  |  |  |
| No | 11(45.83) | 1 |  |  |  |
| Yes | 13(54.17) | 3.47 | 0.004(1.50, 8.0) | 2.78 | 0.04(1.02, 7.51) |
| **Social support** |  |  |  |  |  |
| No | 16(66.7) | 1 |  |  |  |
| Yes | 8(33.3) | 0.46 | 0.08(0.19, 1.12) | 0.45 | 0.117(0.17, 1.21) |
| **Stigma** |  |  |  |  |  |
| No | 19(79.17) | 1 |  |  |  |
| Yes | 5(20.83) | 3.8 | 0.014(1.30, 11.08) | 2.11 | 0.035(1.1, 14.25) |

ART= Anti-retroviral therapy, BMI= Body Mass Index, WHO= World Health Organization, OR= Odds Ratio, AOR= Adjusted Odds Ratio, VF= Virological failure.

### Components of quality of life in relation to gender

Table 4 summarizes the various QoL domains in relation to gender. There were significant relationship between overall QoL (*p=*0.0002), physical domain (*p=*0.005), psychological (*p<*0.0001) and environment domain (*p*<0.0001) in relation to gender. Health satisfaction (*p=*0.9398), level of independence (*p=*0.118), social relationships (*p=*0.1945) and spiritual/religion/personal belief were not significantly related to gender. The overall QoL mean score was 75.35% (SD: 6.77). The physical domain recorded the highest score (86.01%) whilst the environmental domain recorded the least score (62.73%).

**Table 4:** Quality of life components by gender among people with HIV/AIDS at a Municipal Hospital, Volta Region.

| Parameter | Total | Female | Male | *p*-value |
| --- | --- | --- | --- | --- |
| Overall quality of life | 75.35±6.77 | 74.77 | 78.06 | **0.0002** |
| This year health compared to last year | 80.54 | 80.91 | 80.57 | 0.7916 |
| Health satisfaction | 80.65 | 80.61 | 80.86 | 0.9398 |
| Physical | 86.01 | 85.32 | 89.21 | 0.005 |
| Psychological | 78.25 | 77.26 | 82.91 | <0.0001 |
| Level of independence | 71.13 | 70.82 | 72.57 | 0.118 |
| Social relationships | 75.94 | 75.67 | 77.21 | 0.1945 |
| Environment | 62.73 | 61.91 | 66.61 | <0.0001 |
| Spiritual/religion/personal belief | 85.79 | 85.49 | 87.21 | 0.2058 |

### Factors associated with quality of life of people with HIV/AIDS

Table 5 summarizes the association between participants’ characteristics and overall QoL. Variables uch as male gender (*R^2^=*0.048, *P*=0.048) and monthly income (*R^2^*= 2.03, *p=*0.002) were positively ssociated with overall QoL. Also, social support (R^2^=2.94, *p<*0.0001) and comorbidity (R^2^=-2.7, *<0.0001*) were significantly associated with overall QoL.

**Table 5.** Factors associated with quality of life of people with HIV/AIDS at a Municipal Hospital, olta Region.

| Variables | Coefficient ($R^2$) | P-value | CI | |
| --- | --- | --- | --- | --- |
| **Marital status** |  |  |  |  |
| Married | 1.71 | 0.106 | -0.363 | 3.783 |
| Separated | -2.841 | 0.155 | -6.758 | 1.075 |
| Single | 0.959 | 0.533 | -2.061 | 3.981 |
| Widowed | 0.801 | 0.466 | -1.132 | 2.960 |
| **Comorbidity** |  |  |  |  |
| Yes | -2.708 | <0.0001 | -4.209 | -1.207 |
| **Education** |  |  |  |  |
| None | -0.009 | 0.990 | -1.544 | 1.524 |
| Senior High | 0.244 | 0.863 | -2.529 | 5.587 |
| Tertiary | 2.343 | 0.220 | -1.408 | 6.095 |
| **Income (GHS)** |  |  |  |  |
| 500-1599 | 2.037 | 0.002 | 0.729 | 3.345 |
| 1600 Or more | 4.988 | 0.094 | -0.859 | 10.836 |
| **Occupation** |  |  |  |  |
| Self-employed | 2.978 | 0.536 | -2.334 | 0.911 |
| Unemployed | 1.270 | 0.794 | -8.291 | 10.833 |
| **Social support** |  |  |  |  |
| Yes | 3.942 | <0.0001 | 2.696 | 5.187 |

CI= Confidence Interval.

## Discussion

The study determined the prevalence and predictors of virological failure and QoL of PWHA. The prevalence of virological failure observed in this study was 6.03%, which is slightly higher than the global UNAIDS target of 5.0% [31, 32]. The findings of this study, however, is lower when compared with similar studies in Ghana by Abubakari et al. (47%), Owusu et al. (19%) and Boakye et al. (24%) [19–21]. The reason for the lower prevalence observed in this study compared to similar studies in Ghana could be due to introduction of dolutegravir into the care and management of HIV/AIDS in 2019, keeping scheduled appointment, improved social support, HIV disclosure, strict drug adherence and the current asymptomatic state of study participants. The administration of dolutegravir among PWHA has been shown to improve health outcomes, promote high viral suppression rate, has high tolerance rate and higher resistance barrier against mutations [33, 34]. Individuals who disclose their HIV status are more likely to enjoy strong social support ranging from companionship, improved coping strategies, financial support, motivation and emotional support [35, 36]. These support increases their quality of life and promote good ART adherence behaviors, contributing to improved CD4 count, strong immunity and reduced viral load. Also, the current asymptomatic state of study participants suggest that they were responding positively to therapy, thus contributing to their low viral load count.

The study further observed that HIV stigmatization, forgetting to take ART and being classified as baseline WHO staging II among PWHA were associated with virological failure. PWHA who forget to adhere to ART medications were more likely to develop virological failure than those who strictly adhere to medications. Similar observations were made by other researchers [37–39]. Interestingly, most of these individuals who forgot to take their ART medications in this study tend to do so on weekends. This could be partly due to busy weekend schedules, travelling away from home, changes in routine activities and family visit. It was also observed that PWHA who are classified as being in baseline WHO clinical stage II were more likely to develop virological failure. Such individuals are often diagnosed late, and the delay in HIV diagnosis leads to the development of drug resistance [40], opportunistic infections and high viral load. This study finding is consistent with the study in Ethiopia by Desta and colleagues which observed that WHO staging II was associated with the development of virological failure [41]. Another study by Kityo et al. [42] also reported similar findings where WHO staging II contributed to the development of virological failure in Ugandan children. The study further observed that PWHA who reported being stigmatized were more likely to develop virological failure. The observation in this study agrees with a study by Hargreaves and colleagues [43] which also observed that PWHA internalized stigma were associated with virological failure. HIV stigmatization affects interventions such as drug adherence, access to treatment and voluntary counselling and testing.

The overall QoL observed among PWHA in this study was good (75.35%), consistent with a study conducted in Ghana (71.29%) [22] and Indonesia(71.6%) [44]. It is important to emphasize that the environmental domain observed the least QoL score, and is consistent with the findings of other studies conducted in Ghana [45, 46] and Portugal [47]. Monthly income of PWHA was also shown to have contributed to the improved QoL observed in this study. This observation is in agreement with a study by Ebrahimi-Kalan et al. [48] which reported that low monthly income reduces QoL among PWHA in Iran. Similarly, another study by Jiang and colleagues [49] also observed monthly income as a contributor to QoL among men who have sex with other men in China. PWHA who receive high income are able to provide for their health, nutritional and financial needs, and these positively influence their immunity and mental health.

This present study further observed that social support contributes to the overall QoL of PWHA, influencing all the various domains of life assessed on the WHOQOL-HIV BREF tool. Similar findings were made by Abrefa-Gyan and colleagues [50] and Birore et al. [51] where social support positively improved the QoL of PWHA in Ghana. Social support provides financial assistance, improves mental health, promotes HIV disclosure [25] and influences drug adherence among PWHA [52]. It was also noted in this study that, the absence of comorbidities among PWHA contributed to the observed good QoL. These individuals were free from renal impairment, drug to drug interaction and other complications associated with comorbidities. Studies in Ethiopia by Nigusso et al. [16] and Langebeek et al. [53] reported similar findings, attributing improved QoL among PWHA to the absence of comorbidities. It is interesting to note that whereas other studies reported poor QoL among HIV-infected individuals with virological failure, this study observed good QoL among these people.

The use of a cross-sectional study introduced minimal selection bias to the study, however, it enabled efficient data collection within a short time frame, eliminating the need for participant follow-up.

## Conclusion

Virological failure in the municipality was high, exceeding the 5.0% target set by the UNAIDS. Factors such as forgetting to take ART, WHO staging II and HIV stigmatization contributed to the observed virological failure in the municipality. It is therefore necessary that the Government of Ghana in collaboration with the National AIDS Control Programme (NACP) consider implementing the use of long-acting injectable therapy in routine management of HIV/AIDS to improve medication adherence and reduce HIV stigmatization among PWHA. Despite the high virological failure, majority of the study participants presented with good QoL. Factors such as presence of social support, an average monthly income and absence of comorbidities contributed to the improved QoL. Routine assessment of QoL in ART clinics are crucial in improving health outcomes among PWHA. Strengthening social support systems, including peer support groups within ART clinics and broader community networks, could also provide emotional stability and security for PWHA. Also, the NACP should collaborate with financial donors, government agencies and other social welfare organizations to establish employment opportunities and provide both financial and material assistance to economically disadvantaged PWHA.

## Data Availability

All relevant data are within the manuscript and its Supporting Information files.

## Acknowledgement

The authors appreciate the commitment and dedication of participants involved in the study as well as the officials of the Municipality for granting us the permission to undetake this study. We are aslo grateful to Miss. Mercy Demaris Quarm for the technical support provided throughout the study.

---

**Prevalence of virological failure among study participants**

The propo1tion of HIV-infected adults with virological failure is depicted 111 figure 1. The proportion of study participants with virological failure was 6.03% whilst 93.97% had their viral load suppressed.

**Fig 1:**
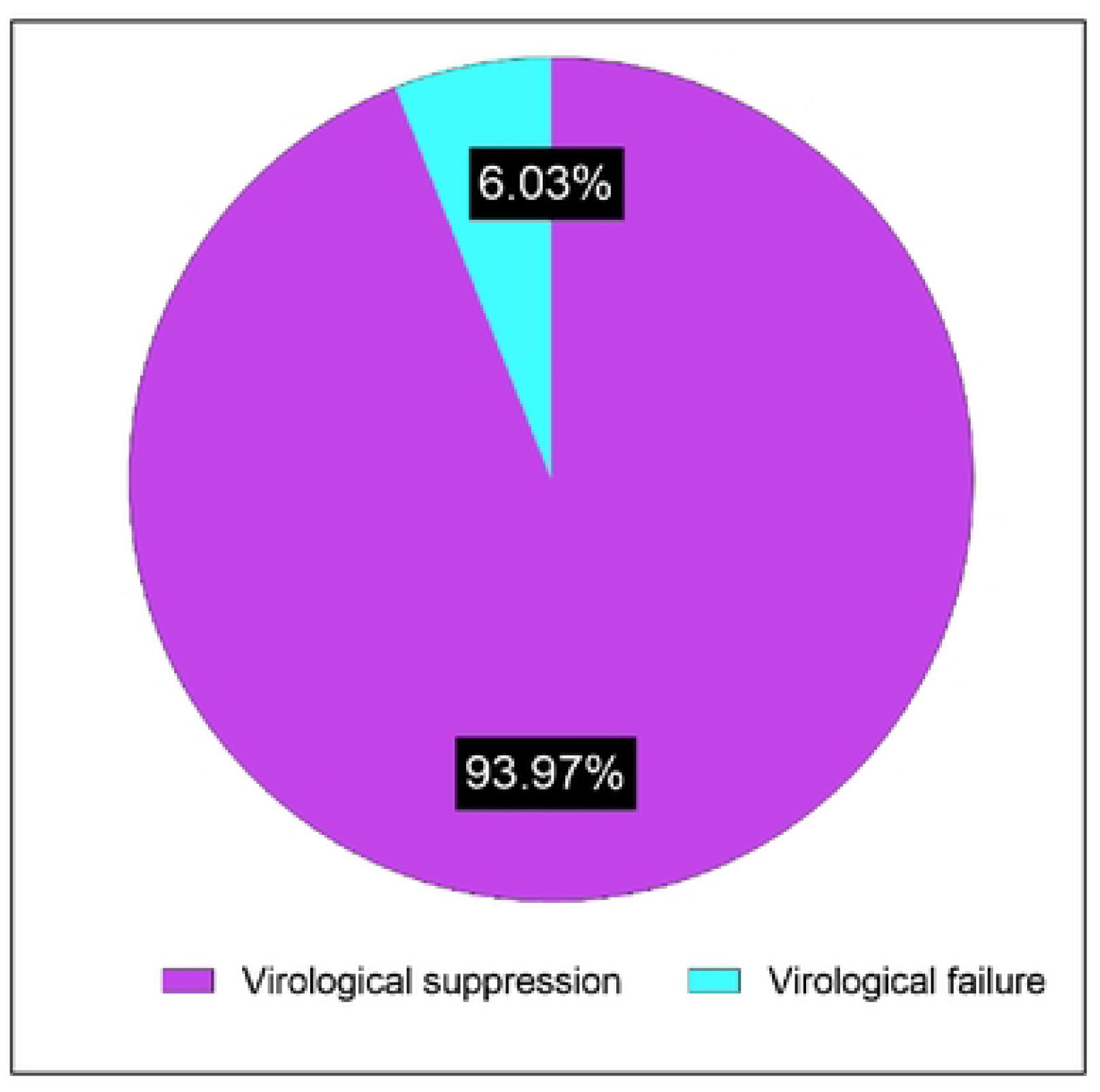
Prevalence of virological failure among HIV-infected adults on ART.

